# Impact of population density on the Covid-19 infected and mortality rates in India

**DOI:** 10.1101/2020.08.21.20179416

**Authors:** Arunava Bhadra, Arindam Mukherjee, Kabita Sarkar

## Abstract

The Covid-19 is a highly contagious disease which becomes a serious global health concern. The residents living in areas with high population density, such as big or metropolitan cities have a higher probability to come into close contact with others and consequently any contagious disease is expected to spread rapidly in dense areas. However, recently after analyzing Covid-19 cases in the US researchers at the Johns Hopkins Bloomberg School of Public Health, London school of economics and IZA – Institute of Labor Economics conclude that the spread of Covid-19 is not linked with population density. Here we investigate the influence of population density on Covid-19 spread and related mortality in the context of India. After a detailed correlation and regression analysis of infection and mortality rates due to Covid-19 at the district level we find moderate association between Covid-19 spread and population density.

## Introduction

The emergence and continuous spreading of the highly contagious disease Covid-19 leads the World in a very distressing stage. The disease has severe adverse effects on the world economy, as well as on many aspects of human lives like employment, education, physical and mental health of individuals, etc.

In the absence of any precise medicine for the treatment of Covid-19 or any effective vaccine to prevent it several efforts are made, based on available pandemic data, in modeling the Covid-19 cases to understand the dynamics of infections (Chen et al., 2020; Roy et al., 2020; Rahaman et al., 2020) and subsequently forecasting about the future course of the pandemic for scheming strategies to quickly contain the spreading of the infections by other means like physical distancing, lockdown, etc.

The modeling of transmission of any infectious disease relies on several factors associated with the disease. A number of studies suggest that Covid-19 infection is associated with meteorological factors such as temperature, humidity, wind speed, etc. In particular, below 3° C the number of COVID-19 infections in China is found to have a positive linear association with average temperature (Zhu & Xie, 2020). A similar correlation between temperature and Covid-19 cases is also found in several Countries and cities, like India (Gupta et al., 2020), Indonesia (Tosepu et al., 2020), Turkey (Sahin, 2020), New York (USA) (Bashir et al., 2020) as well as on a worldwide scale (Chen 2020a).

Since coronavirus (SARS-COV-2) transmits via human contact (Chan et al., 2020; Li et al., 2020) the common perception is that Covid-19 spread rapidly in dense areas whereas the probability of getting infected is low in areas with low population density. However, after analyzing Covid-19 infection and death rates of 913 urban counties in the United States a recent investigation by researchers at the Johns Hopkins Bloomberg School of Public Health claims that the infection rate is not linked with population density whereas death rate is inversely related to population density (Hamidi et al, 2020) except for metropolitan areas where higher infection and higher mortality rates have been noted. The inverse mortality relation with density has been attributed by the authors on the availability of better healthcare systems at higher density locations. A very recent study by the researchers from the London school of economics and IZA – Institute of Labor Economics, based on urban Covid-19 cases in the US concluded that the timing of the outbreak depends on population density with denser regions led an early breakout but both Covid-19 infection and death rates are unrelated with urban population density (Carozzi et al, 2020).

India is a large country with a total population of more than 1.3 billion. It has diverse demographic features. On the other hand as of 10^th^ September 2020, India is the second maximum Covid-19 affected country by the total number of infected people after the USA. The Covid-19 pandemic situation in India thus allows us to cross-check the recent findings (Hamidi et al, 2020; Carozzi et al, 2020) on the impact of population density on Covid-19 spread and related mortality. Probably the first study on the impact of population density (along with several other geographical factors) on the spread of Covid-19 was done by Gupta et al (2020) in the context of India but their analysis was based on the Covid-19 data of Indian states up to the 27^th^ April 2020 when total reported Covid-19 cases was only 29458 and they considered population density at the state level only.

In India, a major part of total infections and mortality have, at least apparently, been attributed to metropolitan cities such as Mumbai, Delhi, Bangalore, Ahmedabad, Chennai, and Kolkata. The infection and death rates appear to be much lower at remote districts. However, because of the large population of the metropolitan cities, the total infections and mortality are normally higher in those places. Moreover, a strong presence of media lead reporting the Covid-19 cases in metropolitan cities in greater detail which might give a false impression that only these cities are dominantly contributing Covid-19 infection and related death.

To have a concrete and reliable idea on the issue in the present work a detailed analysis of Covid-19 infected and death cases as a function of population density of Indian districts are performed. So far there is no established model about the role of population density on the spread of highly contagious diseases. The findings of the impact of population density on mortality during the 1918 influenza pandemic are debatable. A few researchers find no link between population density and death in 1918 influenza (Chowell et al., 2008; Nishiura, & Chowell 2008) whereas a few others claim for a significant positive correlation (Garrett 2010). The Covid-19 pandemic allows us to study the issue in more detail and reliably. The issue is also important in the connection to planning of future towns/cities.

Our findings suggest that till 10^th^ September 2020 Covid-19 infections and mortality in India are moderately correlated with population density; on the average both the infected and the death cases are higher for large population density.

## Data used

India, the seventh-largest country of the world is a federal union of twenty-eight (28) states and eight (8) union territories at present. The states and union territories are further divided into districts for administrative convenience. At present, there are total of 718 districts in India.

The district wise Covid-19 cumulative infection and death cases data were taken from the daily bulletins of the Ministry of Health and Family Welfares of Government of India (https://www.mohfw.gov.in/) and the State Governments (such as https://www.wbhealth.gov.in/pages/corona/bulletin, (West Bengal), http://dgmhup.gov.in/en/covidteamreport, (Uttar Pradesh) https://arogya.maharashtra.gov.in/1175/Novel--Corona-Virus, (Maharashtra) https://stopcorona.tn.gov.in/daily-bulletin/, (Tamil Nadu), http://hmfw.ap.gov.in/covid_19_dailybulletins.aspx (Andhra Pradesh etc.). The daily compiled data from the Central and all the State Government bulletins are also available at api.covid19india.org. We use the data till 10^th^ September, 2020.

The updated corona infected and death data are not available for a few districts such as the districts of Delhi, Goa, Andaman & Nicobars. Besides that, the cumulative infected and mortality data for districts of Telangana are not reported. We, therefore could not include those districts in our analysis.

For the district-wise total population and population density we have used the census 2011 data of the Government of India which are given at https://censusindia.gov.in/. A few new districts have been created since 2011 by splitting, rearranging some districts. For such cases, we have taken the population and density data from the web site of the concerned districts though we could not include all such districts because of lack of data on the district website. We finally consider a total of six hundred districts of India in our analysis.

## Methodology

Our primary objective is to explore whether any correlation between Covid-19 infected and mortality cases in India with population density exists or not and if yes, we shall consider modeling between the Covid-19 infection and mortality rate in India (dependent variables) with population density (independent variable).

The computation of the correlation coefficient is the most commonly adopted approach for judging statistical relationship between two variables which essentially measures the degree of (linear) association (Bewick et al., 2003; Kunter et al., 2005).

For a set of n pair of observations x_i_ and y_i_ (i varies from 1 to n), the (Pearson) correlation coefficient is defined by

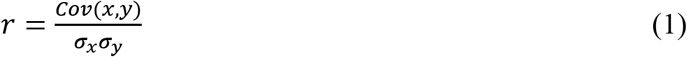

where 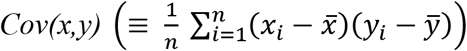 denotes the cross-covariance of the two variables and *σ_i_ (i=x,y)* is the standard deviation.

It is important to examine whether the association is genuine or not which can be done by considering the null hypothesis test. We shall apply the F test, which compares variances of the two variables, and the t-test for the purpose and estimate the p-value which essentially gives the probability that the results from the sample data occurred by chance.

We shall also consider linear regression analysis to model the relationship between the infection/mortality rate and population density. The linear regression relies on the relation

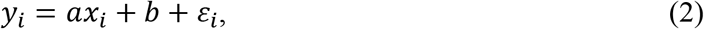

where *x* and *y* are the independent and the dependent variables respectively, *a* is the slope, *b* is the intercept on the *y*-axis and *ε* is the error with zero mean value. In the present work population density is the independent variable, the infection/mortality rate is the dependent variable. Since the infection/mortality rate must vanish when population density is zero, we have opted *b=0*. However, non-zero *b* is also considered for completeness of the analysis which incorporates the option that the behavior of y(x) is non-linear near the origin. Using the method of least squares we estimate the parameters *a^e^* and *b^e^* from the data. So for a given x_j_ the model estimated 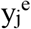 will be

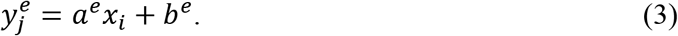

The coefficient of determination (R^2^) quantifies the amount of variability in dependent variable explained by the model and is defined by the relation

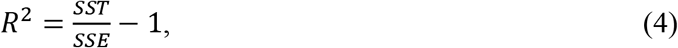

where SSE is the sum of squared errors (squared residuals) 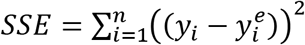, and SST is the sum of squared variation in dependent variable about its mean 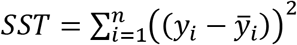. With the increase in number of regressors, the value of R^2^ always increases. For a meaningful measure of goodness of model fit, adjusted R^2^ is used instead of considering the number of explanatory variables in the model. Microsoft MS Excel software has been exploited to compute the mentioned concerned parameters from the data.

## Results and Discussion

The first case of COVID-19 was reported in India on 30th January 2020. By 10^th^ September 2020 the total 4559727 people are infected in India by coronavirus out of which 3539983 have recovered, while it is fatal for 76304 Indians.

Many of the Indian states are quite large and populous. We first address the states separately but restrict our analysis only to scatter diagram. Since the population of different districts are vastly different we have considered the total infected and death cases per one million people for each of the districts. Here we shall show the results for four major states – West Bengal, Maharashtra, Uttar Pradesh, and Tamil Nadu, which are from the East, West, North, and South parts of the country respectively. The scatter plot of the total (cumulative) infected and death cases per one million people against population density for the stated states are depicted in Figure 1. It is found that the infection and death rate are higher in metropolitan cities having huge population density in all the cases except Uttar Pradesh where no districts have very large population density. On the other hand, it is noticed that Covid-19 spread and related death are low in districts with low population density. The situation is not very clear in districts with moderate density; the data points spread all over the graphical area with a slight trend to rise of infection and death with density.

**Figure 1:**
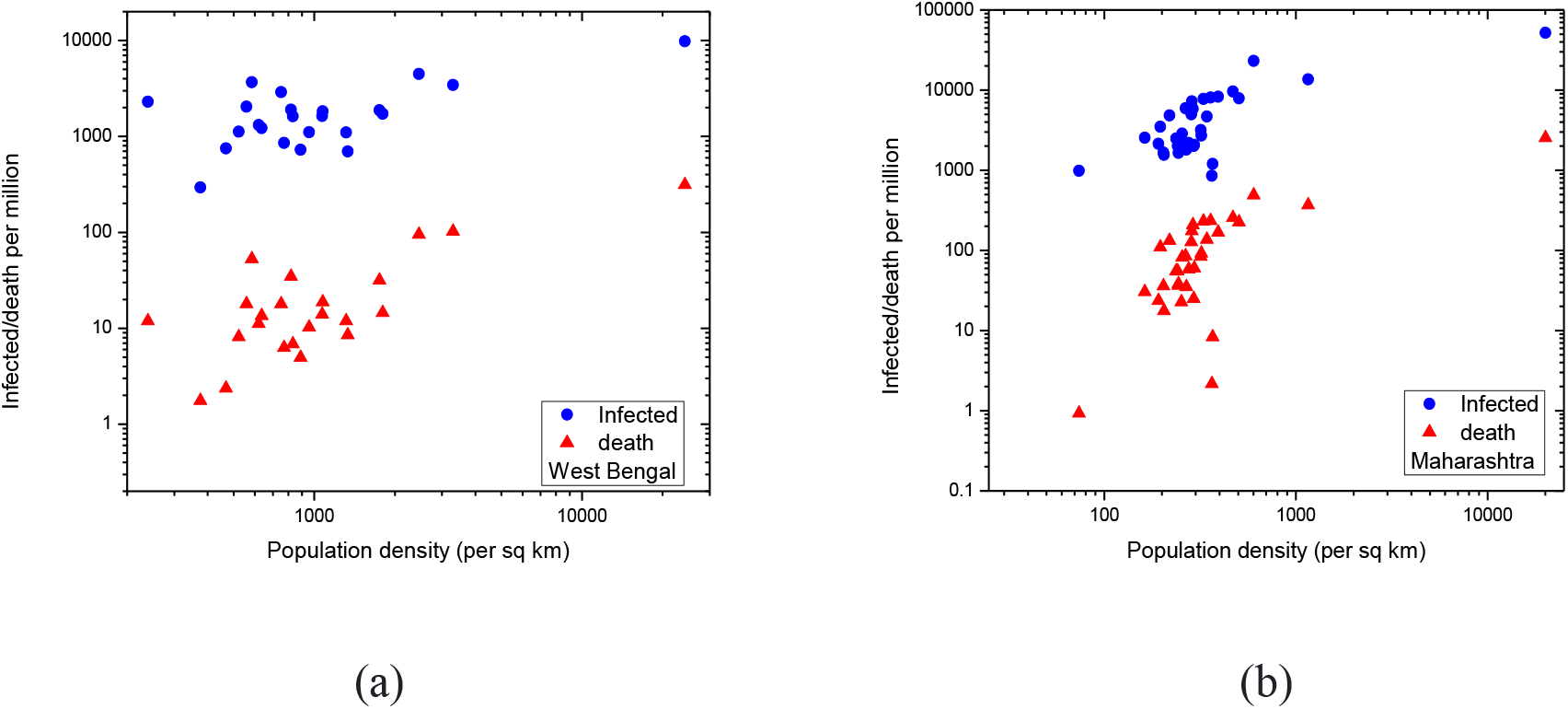
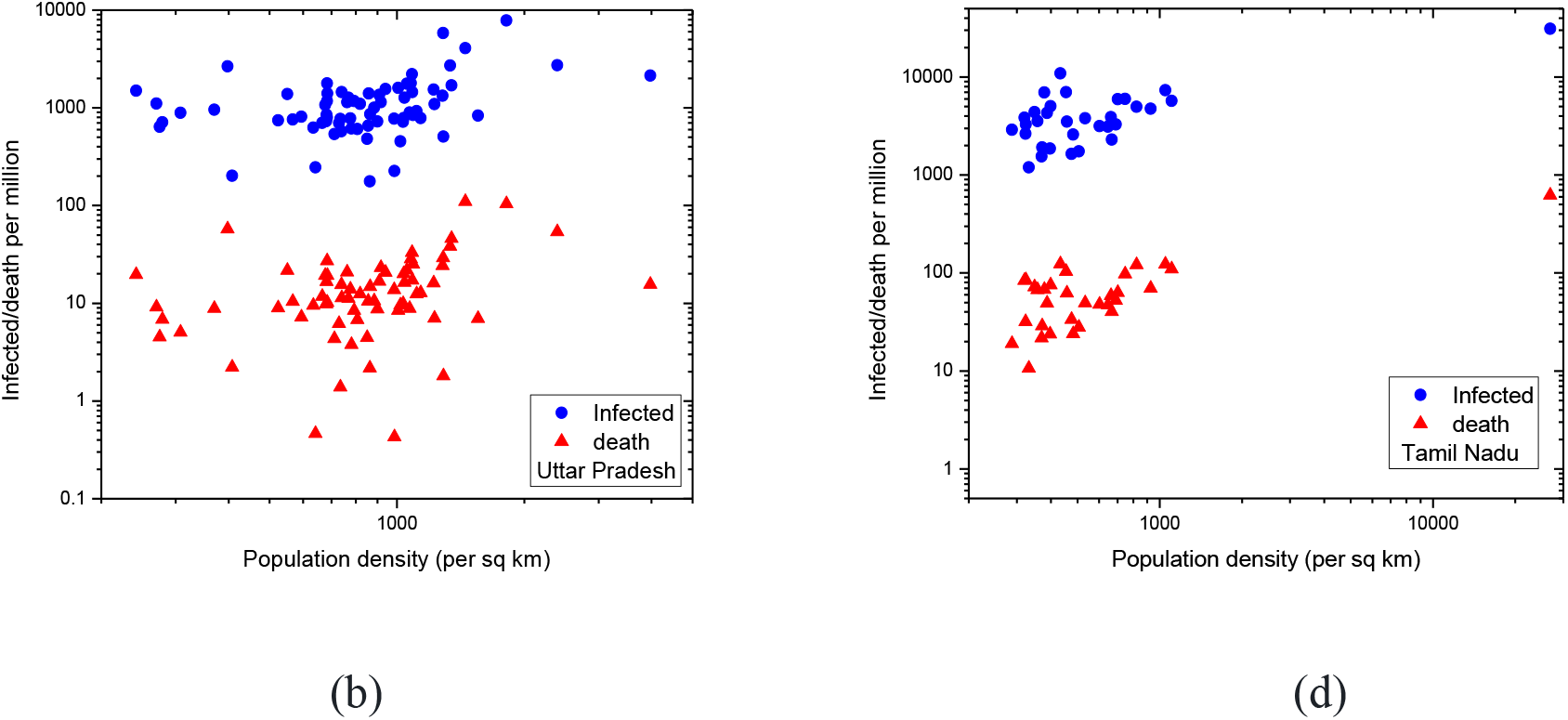
Variation of infected and death rate due to Covid-19 with population density of districts of (a) West Bengal, (b) Maharashtra (C) Uttar Pradesh and (D) Tamil Nadu.

Next we consider India as a whole. The scatter plots of the total (cumulative) infected and death cases per one million people against population density for 600 districts of the country are shown in Figure 2 (a). Though there are large fluctuations, particularly in the mid population density regions, overall there is a tendency that both infection and death cases rise with population density.

**Figure 2:**
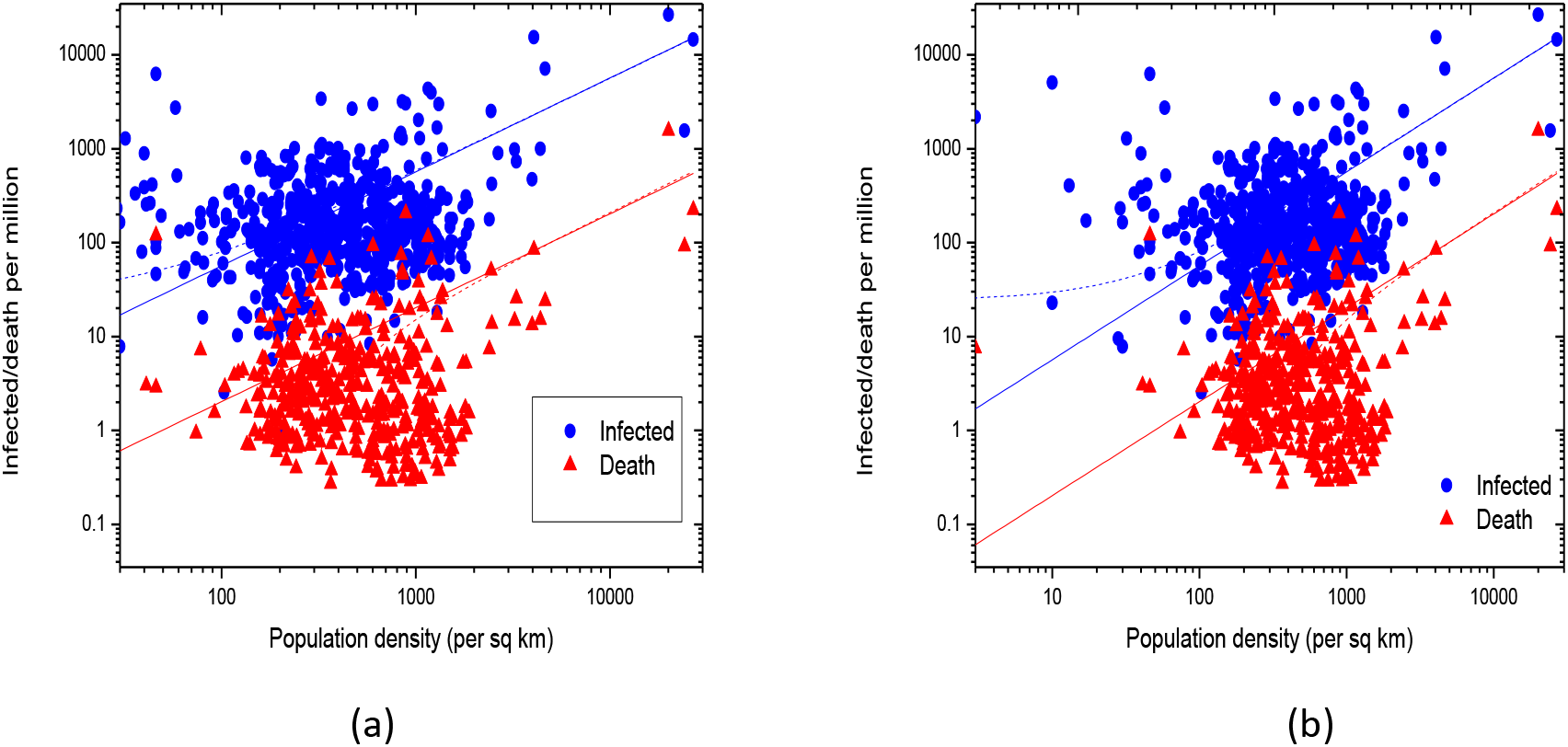
Same as Figure 1 but for the whole India, (a) up to 10^th^ September 2020 (b) till 5^th^ July2020. The solid and dash lines describe the linear fit of the data with zero and non-zero intercepts respectively.

We estimate the correlation coefficients considering infection and mortality rates as the dependent variables and population density as the independent variable which are found as 0.49 (for infection) and 0.59 (mortality) respectively indicating moderate positive correlation. For the significance test, the p-values are computed which are found very small (~10^-37^ for infection rate and even smaller for mortality rate), less than any sensible significant level and hence the null hypothesis (no correlation) is rejected.

Subsequently we express the variation of the infection/mortality rate with population density through equation (3). The least-square fitting gives 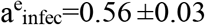 and 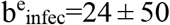 (consistent with no intercept) and 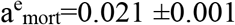 and 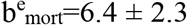. We also force the straight line to pass through the origin since the infection/mortality rate should be zero when population density vanishes. It is found that the correlation coefficients are higher and p-values are lower in no intercept cases. The linear fit of mortality rate against population density passing through the origin gives the adjusted-R^2^ around 41% that decreases to 23% when megacities are excluded in the fitting. It is found that the power-law relation does not describe the data well; it gives much lower R^2^ compare to that of the linear fit.

The correlation between population density and infected/death cases is revealed when we consider small bins in population density and corresponding average infection/death cases are taken. In the dataset of districts of India, the population density per square km is found to vary from 3 to 26903. We bin the population density in log scale with varying widths; particularly larger bin widths at high population density are considered so as to have at least a reasonable count of districts (at least 3) in each bin. The results are shown in Figure 3. A linear (proportional) fit describes both the infection and mortality data well.

**Figure 3.**
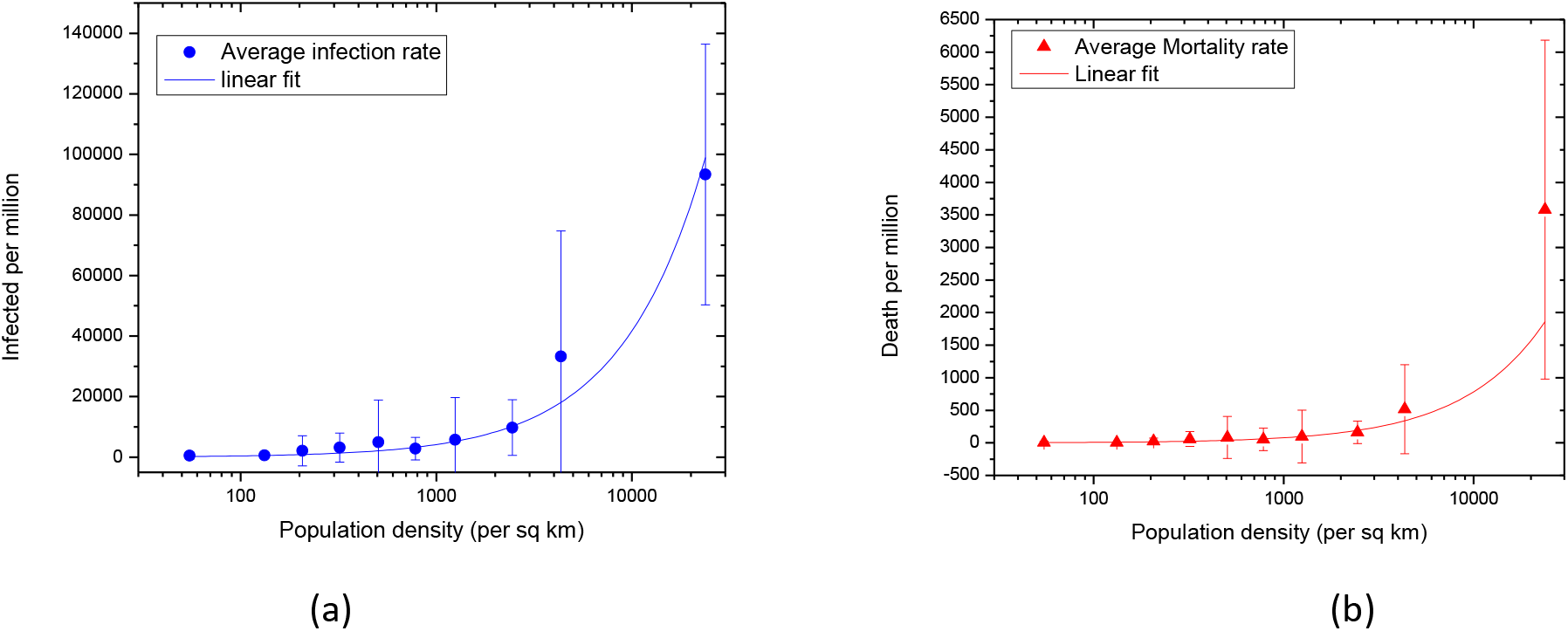
Variation of averaged infected and death rate due to Covid-19 with population density for the whole country.

To verify the claim of researchers that denser regions led an early breakout we plot the same Covid-19 infection/death variation with population density in figure 2(b) but for the data till 5^th^ July 2020. It appears that with the time the number of infection and death cases are increased but the overall trend remains mostly unaltered at least till 10^th^ September, 2020.

**Table 1:** Correlation coefficients and p-values

| Data | Infection |  |  |  | mortality |  |  |  |
| --- | --- | --- | --- | --- | --- | --- | --- | --- |
|  | Linear fit |  | Linear fit with no intercept |  | Linear fit |  | Linear fit with no intercept |  |
|  | r | p | r | p | r | p | r | p |
| Up to 10<sup>th</sup> September 2020 | 0.49 | 0.0000 | 0.58 | 0.0000 | 0.59 | 0.0000 | 0.64 | 0.0000 |
| Up to 5<sup>th</sup> July 2020 | 0.65 | 0.0000 | 0.67 | 0.0000 | 0.57 | 0.0000 | 0.57 | 0.0000 |

**Table 2:** Adjusted R^2^ in Linear fitting of Infection and death rate.

| Data | Infection | mortality |
| --- | --- | --- |

|  | Linear fit | Linear fit with no intercept | Linear fit | Linear fit with no intercept |
| --- | --- | --- | --- | --- |
| Up to 10<sup>th</sup> September 2020 | 0.24 | 0.33 | 0.35 | 0.41 |
| Up to 5<sup>th</sup> July 2020 | 0.42 | 0.45 | 0.32 | 0.32 |
| Up to 10<sup>th</sup> September 2020 but excluding the megacities | 0.11 | 0.38 | 0.32 | 0.23 |
| Up to 5<sup>th</sup> July 2020 but excluding the megacities | 0.11 | 0.22 | 0.03 | 0.12 |

In the present analysis we have considered population density obtained dividing the total population of a district by its total area which implicitly assumes a uniform spatial distribution of populations within a district. The Covid-19 spread and death cases are also considered as uniform. Such assumptions lead to some uncertainty in estimating the correlation between Covid-19 spread and population density. For instance, the population density in the Darjeeling district of West Bengal is among the lowest in the state though the infection and mortality cases are relatively high in the district. This is because of Siliguri, which is one of the congested cities of the state, falls in the Darjeeling district. Most of the infection and mortality cases in the Darjeeling district (and also in the Jalpaiguri district which jurisdiction wise contain a part of Siliguri) are from Siliguri. Whereas the major geographical part of the Darjeeling district is the hill area where population density is quite low and thereby the resulting population density of the district is on the lower side. Further study is needed to overcome the stated issues, say by considering the weighted population mean which is a measure of average “experienced” density or considering sub-division as a basic unit for the study. However, the limited availability of necessary data in the public domain is a major hindrance to such improvements.

The R^2^ value of the relation between infection/mortality rate and population density is found moderate, not very high, which implies that only a part of the infection/mortality rate due to Covid-19 can be explained in terms of population density. This can be understood by the facts that the Covid-19 spread and related mortality in a district may depend on various other factors including geographical features, economic conditions, prevailing health condition**s**, genetic factors, health infrastructure, policies adopted by the regulating authorities, the average age of the residents of the districts, number of testing, etc. It is already known that Covid-19 has a larger impact on the older population (and population with comorbidity). The share of the older population is not the same in all the districts, neither the total testing number. However, there is no prevailing straightforward and unambiguous method of entangling different probable dependent factors. More research on these aspects are necessary to address the stated issues.

## Conclusion

In conclusion the present analysis indicates a positive correlation between Covid-19 infection and related mortality with population density as revealed from the correlation and R^2^ analysis, in contrast to the findings by the researchers at the Johns Hopkins Bloomberg School of Public Health and London school of economics and IZA – Institute of Labor Economics, based on US data. The significance of the correlation is found high from the p-values which strongly indicate that the null hypothesis is false in the present case, at least till now.

There is a vast difference in the living conditions of people in the US and in India which may be responsible for the different behavior of the infected/mortality cases due to Covid-19 with population density in the two countries. The (large) density in India is reflected through the pressing of people against each other in the street, public vehicles, trains, queue for ration, etc. The average area occupied by a family for living in cities of India is also much smaller. The Covid-19 cases were found higher in metropolitan cities even in the USA (Hamidi et al, 2020). So containing a highly infectious disease like Covid-19 is a serious challenge for the country like India unless by any means (maybe by mutation also) the coronavirus (SARS-COV-2) is transformed to a non-lethal variety or an effective vaccine/medicine is discovered.

## Data Availability

The district wise Covid-19 cumulative infection and death cases data for India are taken from the daily bulletins of the Ministry of Health and Family Welfares of Government of India (https://www.mohfw.gov.in/) and the State Governments as compiled also at api.covid19india.org

## Compliance with ethical standards

### Conflict of interest

There is no conflict of interest between the authors.

## Notes

### Competing Interest Statement

The authors have declared no competing interest.

### Funding Statement

No support.

### Summary of Updates

The methodology is described. The data of the analysis is updated.

## References

Bewick, V., Cheek, L., and Ball, J. (2003) Statistics review 7: Correlation and regression, Critical Care 7:451–459 (DOI 10.1186/cc2401)

Carozzi, F., Provenzano, S. and Roth, S. (2020) Urban Density and COVID-19, Discussion Paper Series, IZA Institure of Labor Economics, IZA DP No. 13440

Chan, J.F.-W., Yuan, S., Kok, K.-H., To, K.K.-W., Chu, H., Yang, J., Xing, F., … Yuen, K.-Y., (2020). A familial cluster of pneumonia associated with the 2019 novel coronavirus indicating person-to-person transmission: a study of a family cluster. Lancet 395, 514–523. https://doi.org/10.1016/S0140-6736(20)30154-9.

Chen, T., Rui, J., Wang, Q., Zhao, Z., Cui J., Yin, L., (2020), A mathematical model for simulating the phase-based transmissibility of a novel coronavirus., Infectious Diseases of Poverty, 9, 24.

Chen, B., Liang, H., Yuan, X., Hu, Y., Xu, M., Zhao, Y., Zhang, B., Tian, F., and Zhu, X., (2020a). Roles of meteorological conditions in COVID-19 transmission on a worldwide scale. medRxiv https://doi.org/10.1101/2020.03.16.20037168.

Chowell, G., Bettencourt, L. M., Johnson, N., Alonso, W. J., & Viboud, C. (2008). The 1918–1919 influenza pandemic in England and Wales: Spatial patterns in transmissibility and mortality impact. Proceedings of the Royal Society B: Biological Sciences, 275: 501–509. https://doi.org/10.1098/rspb.2007.1477

Garrett, T. A. (2010). Economic effects of the 1918 influenza pandemic: Implications for a modern-day pandemic. Working paper CA0721.2007. Federal Reserve Bank of St. Louis. https://www.stlouisfed.org/_/media/files/pdfs/community-development/research-reports/pandemic_flu_report.pdf

Gupta, A., Banerjee, S. & Das, S. (2020) Significance of geographical factors to the COVID-19 outbreak in India. Model. Earth Syst. Environ.. https://doi.org/10.1007/s40808-020-00838-2

Hamidi, S. Sabouri, S. & Ewing, R. (2020): Does Density Aggravate the COVID-19 Pandemic?, Journal of the American Planning Association, https://doi.org/10.1080/01944363.2020.1777891

Kutner, M. H., Nachtsheim, C. J., Neter, J., Li, W., (2005) Applied Linear Statistical Models, 5^th^ edn. McGraw-Hill/Irwin

Li, Q., Guan, X., Wu, P.,Wang, X., Zhou, L., Tong, Y., … Feng, Z., (2020). Early transmission dynamics in Wuhan, China, of novel coronavirus–infected pneumonia. N. Engl. J. Med. 382: 1199–1207. https://doi.org/10.1056/NEJMoa2001316.

Nishiura, H., & Chowell, G. (2008). Rurality and pandemic influenza: Geographic heterogeneity in the risks of infection and death in Kanagawa, Japan (1918–1919). The New Zealand Medical Journal, 121: 18–27

Rahman, M.R., Islam, A.H.M.H. & Islam, M.N. (2020) Geospatial modelling on the spread and dynamics of 154 day outbreak of the novel coronavirus (COVID-19) pandemic in Bangladesh towards vulnerability zoning and management approaches. Model. Earth Syst. Environ.. https://doi.org/10.1007/s40808-020-00962-z

Roy, S., Bhunia, G.S. & Shit, P.K. (2020) Spatial prediction of COVID-19 epidemic using ARIMA techniques in India. Model. Earth Syst. Environ.. https://doi.org/10.1007/s40808-020-00890-y,

Zhu, Y., Xie, J., (2020). Association between ambient temperature and COVID-19 infection in 122 cities from China. Sci. Total Environ, 724: 138201 https://doi.org/10.1016/j.scitotenv.2020.138201

